# Open label phase I/II clinical trial and predicted efficacy of SARS-CoV-2 RBD protein vaccines SOBERANA 02 and SOBERANA Plus in children

**DOI:** 10.1101/2022.03.03.22271313

**Authors:** Rinaldo Puga-Gómez, Yariset Ricardo-Delgado, Chaumey Rojas-Iriarte, Leyanis Céspedes-Henriquez, Misleidys Piedra-Bello, Dania Vega-Mendoza, Noelvia Pestana Pérez, Beatriz Paredes-Moreno, Meiby Rodríguez-González, Carmen Valenzuela-Silva, Belinda Sánchez-Ramírez, Laura Rodríguez-Noda, Rocmira Pérez-Nicado, Raul González-Mugica, Tays Hernández-García, Talía Fundora-Barrios, Martha Dubet Echevarría, Juliet María Enriquez-Puertas, Yenicet Infante Hernández, Ariel Palenzuela-Díaz, Evelyn Gato-Orozco, Yanet Chappi-Estévez, Julio Cesar Francisco-Pérez, Miladi Suarez Martinez, Ismavy C. Castillo-Quintana, Sonsire Fernandez-Castillo, Yanet Climent-Ruiz, Darielys Santana-Mederos, Yanelda García-Vega, María Eugenia Toledo-Romani, Delaram Doroud, Alireza Biglari, Yury Valdés-Balbín, Dagmar García-Rivera, Vicente Vérez-Bencomo, SOBERANA Research Group

## Abstract

**Objectives:** To evaluate heterologous vaccination scheme in children 3-18 y/o combining two SARS-CoV-2 r-RBD protein vaccines.

**Methods:** A phase I/II open-label, adaptive and multicenter trial evaluated the safety and immunogenicity of two doses of SOBERANA02 and a heterologous third dose of SOBERANA Plus in 350 children 3-18 y/o in Havana Cuba. Primary outcomes were safety (in phase I) and safety/immunogenicity (in phase II) measured by anti-RBD IgG ELISA, molecular and live-virus neutralization titers and specific T-cells response. A comparison with adult‘s immunogenicity and prediction of efficacy were done based on immunological results

**Results:** Local pain was the unique adverse event with frequency >10%, none was serious or severe. Two doses of SOBERANA 02 elicited humoral immune response similar to natural infection; the third dose increased significantly the response in all children, similar to that achieved in vaccinated young adults and higher than in convalescents children. The neutralizing titer was evaluated in a participant‘s subset: GMT was 173.8 (CI 95% 131.7; 229.5) vs. alpha, 142 (CI 95% 101.3; 198.9) vs. delta and 24.8 (CI 95% 16.8; 36.6) vs. beta. An efficacy > 90% was estimated.

**Conclusion:** The heterologous scheme was safe and immunogenic in children 3-18 y/o. Trial registry: https://rpcec.sld.cu/trials/RPCEC00000374

## Introduction

Children protection against COVID-19 is pivotal for controlling virus dissemination and reducing disease incidence. COVID-19 cases and hospitalizations among children and adolescents, firstly driven by the delta variant and recently by omicron, have risen sharply even in countries with high adult vaccination coverage (Delahoy et al., 2021; Elliott et al., 2021). This context has accelerated the clinical trials of anti-SARS-CoV-2 vaccines for children (Xia et al., 2021; Han et al., 2021; Ali et al., 202; Frenck et al., 2021; Walter et al., 2021; Wallace et al., 2021).

For more than 30 years the Finlay Vaccine Institute has produced tetanus toxoid-conjugated vaccines, applied to children worldwide; their safety has been extensively proved through hundreds of millions of doses (Verez-Bencomo et al., 2004; Huang and Wu., 2010). SOBERANA 02 immunogen is anti SARS-CoV-2 recombinant RBD conjugated to tetanus toxoid (Valdes-Balbin et al., 2021a; 2021b). It is the unique conjugate vaccine in WHO‘s vaccine pipeline (WHO COVID-19, 2021). T-cell epitopes present in tetanus toxoid were expected to promote RBD specific B- and T-cell memory, and high affinity, longstanding RBD IgG antibodies.

SOBERANA 02 has proved its safety and immunogenicity in adults 19-80 y/o; after two doses, its efficacy was 71%. Combined with a third dose of SOBERANA Plus (recombinant RBD dimer vaccine) in a three-dose heterologous scheme, efficacy increased to 92.4% (Toledo-Romani et al., 2021a; 2021b; 2021c). On August 2021, the Cuban Regulatory Authority granted their emergency use authorization in adults, being since then extensively applied nationally for preventing COVID-19 in Cuba (CECMED, 2021).

Here, we report the results of an open label phase I/II clinical trial in children 3-18 y/o to evaluate the safety and immunogenicity of two doses of SOBERANA 02 and a third dose of SOBERANA Plus. We avoided a placebo-controlled trial in this age group due to ethical concerns (Dal-Ré and Caplan., 2021); alternatively, a recommended comparison (or immunobridging) with adult‘s immunogenicity was established (FDA, 2021) and the clinical efficacy was estimated based on immunological results.

## Method

### Study design

We designed a phase I/II study, open-label, adaptive and multicenter to evaluate the safety, reactogenicity and immunogenicity of two doses of SOBERANA02 and a heterologous third dose of SOBERANA Plus in children (3-11 y/o) and teenager (12-18 y/o). Two interim analyses would decide interruption/continuation of the study, depending on serious adverse events (AEs) during phase I.

Phase I was conceived in two-step, incorporating firstly 25 children 12-18 y/o (sequence 1). The first interim report (no serious AE detected) seven days after their vaccination allowed incorporating 25 children aged 3-11 (phase 1, sequence 2 and starting phase II in 12-18 y/o (n=150). A second interim report seven days after sequence 2 (no serious AE detected) allowed starting phase II in children 3-11 y/o (n=150). (Figure 1). Detailed information on trial sites is presented in Supplementary Material I

**Figure 1.**
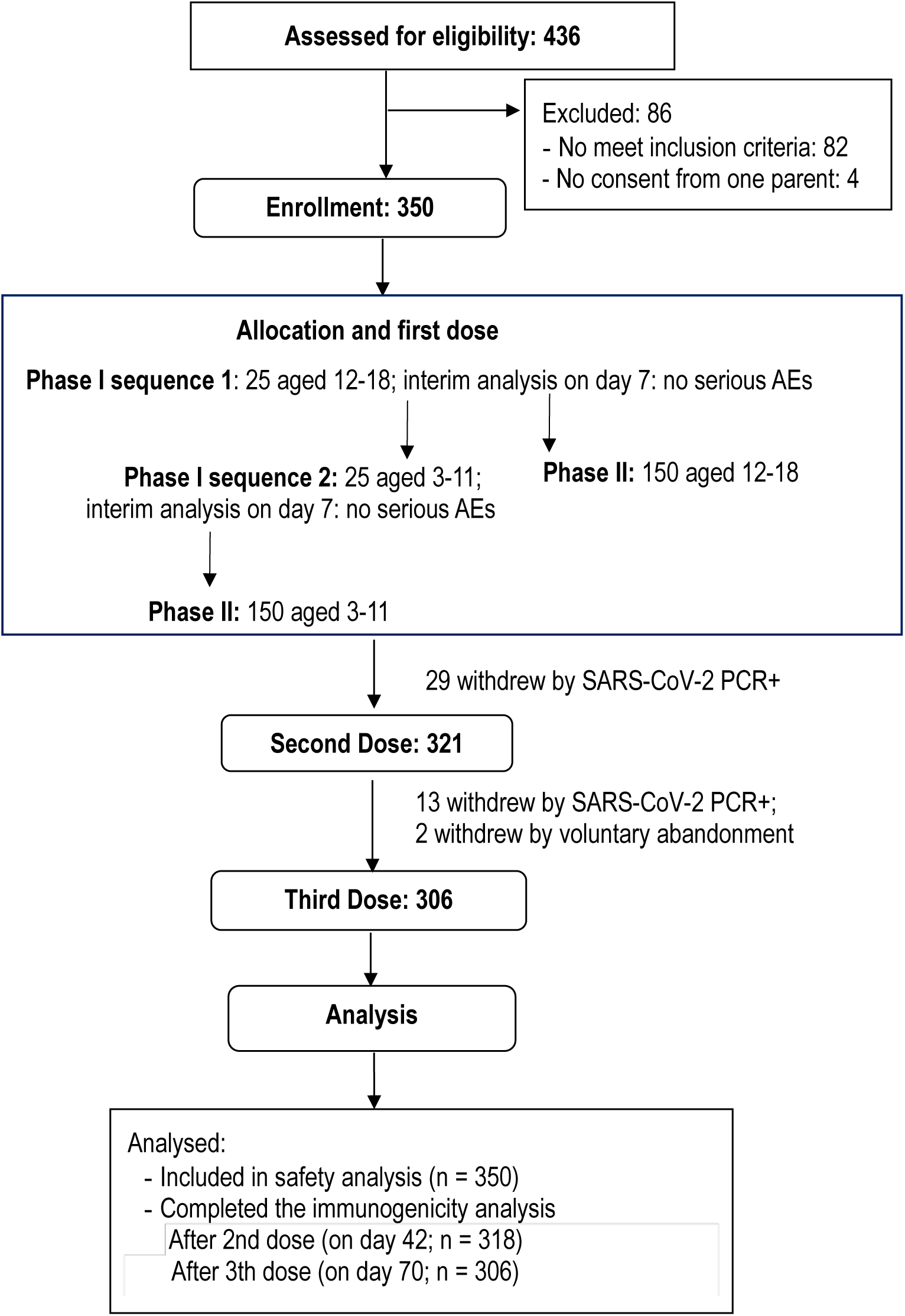
**Flow chart: recruitment, inclusion, vaccination and follow up of 3-18 years old children in phase I/II trial.**

Children were recruited at community level across the primary health system by medical doctors. They were included following physical examination, parent interview and, for phase I, clinical laboratory assays. Key inclusion criteria were weight-height nutritional assessment; physical examination without alterations; clinical laboratory results within the range of reference values (only phase I) and microbiology laboratory tests. Key exclusion criteria were any acute infection, previous or current history of SARS CoV-2 infection and being a contact of a positive COVID-19 case. A detailed description of selection criteria appears in Supplementary Material II.

### Ethical issues

The trial was approved by the Ethical Committee at the “Juan Manuel Marquez” Pediatric Hospital and endorsed by the Cuban National Pediatric Group. The Cuban National Regulatory Agency (Centre for State Control of Medicines and Medical Devices, CECMED) approved the trial (June10^th^, 2021, Authorization Reference: 05.010.21BA).

Independent Data Monitoring Committees formed by five external members specialized on pediatric clinical practice; immunology and statistics were in charge of two interim analyses during phase I.

The trial was conducted according to Helsinki’s Declaration, Good Clinical Practice and the Cuban National Immunization Program. During recruitment, the medical investigators provided to the parents, both orally and written, all information about the vaccine and potential risks and benefits. Written informed consent was obtained from both parents; children ≥over 12 y/o should assent. The decision to participate was not remunerated.

The National Clinical Trials Coordinating Centre (CENCEC) was responsible for monitoring the trial in terms of adherence to the protocol, to Good Clinical Practice and data accuracy.

Trial registry: RPCEC00000374 (Cuban Public Registry of Clinical Trials and WHO International Clinical Registry Trials Platform) (IRCT, 2021).

### Products under evaluation

SOBERANA 02 (RBD chemically conjugated to the carrier protein tetanus toxoid) and SOBERANA Plus (RBD dimer), adjuvated in alumina hydroxide were produced at the Finlay Vaccine Institute and the Centre for Molecular Immunology, in Havana, Cuba, under GMP conditions. Both are subunit vaccines based SARS-CoV-2 RBD, sequence Arg319-Phe541 produced in genetically modified CHO cells. Formulations are detailed in Supplementary Material III-Table S1.

Product batches used: SOBERANA 02 (E1002S02X, E1002S02); SOBERANA Plus (E1001SP).

### Procedures

RT-PCR SARS-CoV-2 was performed to all participants at least 72 hours before each dose. Participants with negative PCR results received the vaccine by intramuscular injections in the deltoid region.

#### Immunization schedule

two doses of SOBERANA 02 and a heterologous third dose of SOBERANA Plus 28 days apart (immunization on days 0, 28, 56). After each immunization, participants were on-site evaluated during one hour. Medical controls visits were planned at 24, 48 and 72 hours, and on days 7, 14 and 28 after each dose. AEs were parents-registered daily. Serum samples were collected on day 0 (before vaccination) and 14 days after the second and the third dose (days 42 and 70). Peripheral blood mononuclear cells were obtained before vaccination and after the third dose (day 70) in a participants’ subset of 45 children randomly selected in each age subgroup.

### Outcomes

Primary outcomes. Phase I: occurrence of serious AEs, measured daily during 28 days after each dose. Phase II: % of subjects with seroconversion ≥4 fold increase of IgG anti-RBD over pre-immunization, on days 42 and 70.

Secondary outcomes. Phase I and phase II: Solicited local and systemic AEs, measured during 7 days after each dose; unsolicited AEs, measured during 28 days after each dose; neutralizing antibody titers (on days 42 and 70, on a sample subset), inhibition of RBD-hACE2 interaction (on days 42, 70). Phase II: Occurrence of serious AEs, measured during 28 days after each dose. Outcomes are detailed in Supplementary Material III).

Outcomes and safety assessment are detailed in Supplementary Material IV and V.

### Immunogenicity assessment

Immunogenicity was evaluated by: a) quantitative ultramicro ELISA (UMELISA SARS-CoV-2 anti-RBD; b) competitive ELISA determined the inhibitory capacity of antibodies for blocking the RBD-hACE2 interaction, expressed as % inhibition and molecular virus neutralization titer (mVNT_50_); c) conventional virus neutralization titer (cVNT_50_) vs. D614G, alpha, beta, and delta variants; d) RBD-specific T-cell response producing IFN-γ and IL-4. Immunogenicity assessment and techniques are described in Supplementary Material VI. All immunological evaluations were performed by external laboratories from the Centre for Immunoassays, Centre of Molecular Immunology and National Civil Defense Research Laboratory. T cell response was evaluated at Finlay Vaccine Institute. A detailed description of immunogenicity assessment and techniques are described in Supplementary Material VI.

### Children Convalescent Serum Panel

A Cuban children convalescent serum panel (CCCSP) was made with sera from 82 patients (3-18 y/o) recovered from COVID-19. Detailed information about panel composition and immunocharacterization is presented in Supplementary Material VII

## Statistical analysis

For phase I, the calculation of sample size was done considering a two-sided 95% confidence interval for one proportion with a width equal to 0.09 to estimate a serious AE rate <1%. For phase II a similar method was used to estimate a seroconversion around 50%, with a lower bound of the confidence interval >30% (trial hypothesis) and a dropout of 20%. This resulted in a sample size of 350 subjects (including subjects from phase I). Detailed statistical tools, procedures and definitions are presented in Supplementary Material VIII.

## Results

Figure 1 and Table 1 describe the study design and demographic characteristics of the participants. From 11^th^ June to 14^th^ July 2021, 426 children (3-18 y/o) were recruited, 350 that accomplished selection criteria were included, and 306 completed the study. There was a balanced ratio on sex and ethnicity; mean age was 11.3 years (SD 4.5).

**Table 1:** Demographic characteristics of subjects included in the clinical trial.

|  | Age groups |  |  |
| --- | --- | --- | --- |
|  | 3-11 years | 12-18 years | Total |
| **N** | 175 | 175 | 350 |
| **Sex** |  |  |  |
| Female | 80 (45.7%) | 83 (47.4%) | 163 (46.6%) |
| Male | 95 (54.3%) | 92 (52.6%) | 187 (53.4%) |
| **Skin colour** |  |  |  |
| White | 122 (69.7%) | 116 (66.3%) | 238 (68.0%) |
| Black | 9 (5.1%) | 11 (6.3%) | 20 (5.7%) |
| Multiracial | 44 (25.1%) | 48 (27.4%) | 92 (26.3%) |
| **Age (years)** |  |  |  |
| Mean (SD) | 7.4 (2.5) | 15.1 (2.1) | 11.3 ± 4.5 |
| Median (IQR) | 8.0 (5.0) | 15.0 (4.0) | 11.5 ± 7.0 |
| Range | 3; 11 | 12;18 | 3-18 |
| **Weight (kg)** |  |  |  |
| Mean (SD) | 29.4 (10.1) | 54.7 (9.0) | 42.0 ± 15.9 |
| Median (IQR) | 27.5 (14.0) | 55.0 (13.0) | 43.0 ± 27.7 |
| Range | 13.0; 58.0 | 32.0; 80.0 | 13.0; 80.0 |
| **Height (cm)** |  |  |  |
| Mean (SD) | 129.1 (17.2) | 164.3 (9.6) | 146.7 ± 22.5 |
| Median (IQR) | 131.0 (26.0) | 164.0 (13.0) | 151.0 ± 34.0 |
| Range | 92; 172 | 142; 190 | 92-190 |
| **BMI (kg/m<sup>2</sup>)** |  |  |  |
| Mean (SD) | 17.0 (2.0) | 20.2 (2.3) | 18.6 ± 2.7 |
| Median (IQR) | 16.7 (2.7) | 19.9 (3.8) | 18.3 ± 4.1 |
| Range | 13.2; 22.8 | 14.6; 25.5 | 13.2-25.5 |
| Data are n (%) unless otherwise specified. Mean (SD)=Mean ± Standard Deviation. Median (IQR)=Median ± Interquartile Range. BMI=Body mass index. Range=(Minimum; Maximum) |  |  |  |

Phase I started by vaccinating 25 children 12-18 y/o with SOBERANA 02; the first interim analysis done seven days after vaccination indicated absence of serious AEs. In consequence, the trial proceeded to phase I, sequence 2, incorporating 25 children aged 3-11 and 150 children aged 12-18 of phase II. The second interim analysis showed no serious AE in children 3-11 y/o (sequence 2); the trial completed phase II vaccinating 150 children 3-11 y/o with SOBERANA 02 first dose.

During the vaccination scheme, 86 children (53.1%) suffered at least one AE; frequency was higher (60%) in teenagers than in young children (46.3%). Severe and serious vaccine-associated AEs (VAAE) did not occur (Table 2). Local AE predominated; the most common was local pain (47.7%), all others had frequencies <5%; only 1.1% reported fever. More than 90% of AEs were classified as mild and lasted ≤72 hours, and 88.5% were associated with vaccination (Table 3, Supplemental Material IX-Table S2). AEs were more frequent after the first dose than after the second and the third dose (Supplemental Material IX-Table S3). Few unsolicited adverse events were recorded (Supplemental Material V-Table S4). No clinically relevant changes were observed in hematology and blood chemistry analyses (Supplemental Material IX-Table S5).

**Table 2:** General characteristics of adverse events.

|  | Age groups |  |  |
| --- | --- | --- | --- |
|  | 3-11 years | 12-18 years | Total |
| **N** | 175 | 175 | 350 |
| Subjects with some AE | 81 (46.3%) | 105 (60.0%) | 186 (53.10%) |
| Subjects with some VAAE | 76 (43.4%) | 101 (57.7%) | 177 (50.6%) |
| Subjects with some Serious AE | 0 (0.0%) | 1 (0.6%) | 1 (0.3%) |
| Subjects with some Serious VAAE | 0 (0.0%) | 0 (0.0%) | 0 (0.0%) |
| Subjects with some Severe AE | 0 (0.0%) | 0 (0.0%) | 0 (0.0%) |
| Subjects with some Severe VAAE | 0 (0.0%) | 0 (0.0%) | 0 (0.0%) |
| **Total Adverse Events** | 141 | 182 | 323 |
| Mild AE | 135 (95.7%) | 167 (91.8%) | 302 (93.5%) |
| Moderate AE | 6 (4.3%) | 15 (8.2%) | 21 (6.5%) |
| Severe AE | 0 (0.0%) | 0 (0.0%) | 0 (0.0%) |
| **Serious AE** | 0 (0.0%) | 1 (0.5%)* | 1 (0.3%) * |
| **Local AE** | 118 (83.7%) | 147 (80.8%) | 265 (82.0%) |
| **Systemic AE** | 23 (16.3%) | 35 (19.2%) | 58 (18.0%) |
| **VAAE** | 126 (89.4%) | 160 (87.9) | 286 (88.5%) |
| **Serious VAAE** | 0 (0.0%) | 0 (0.0%) | 0 (0.0%) |
| **Severe VAAE** | 0 (0.0%) | 0 (0.0%) | 0 (0.0%) |

Data are n (%). AE=Adverse Event. VAAE=Vaccine-Associated Adverse Event; \* Serious AE: Dengue required hospitalization.

**Table 3:** Frequency of solicited adverse events.

|  | Age groups |  |  |
| --- | --- | --- | --- |
|  | 3-11 years | 12-18 years | Total |
| **N** | 175 | 175 | 350 |
| **Subjects with some AE** | 81 (46.3%) | 105 (60.0%) | 186 (53.10%) |
| **Subjects with solicited local AE** |  |  |  |
| Any | 74 (42.3%) | 98 (56.0%) | 172 (49.1%) |
| Local pain | 69 (39.4%) | 98 (56.0%) | 167 (47.7%) |
| Swelling | 9 (5.1%) | 2 (1.1%) | 11 (3.1%) |
| Local warm | 4 (2.3%) | 0 (0.0%) | 4 (1.1%) |
| Erythema | 5 (2.9%) | 1 (0.6%) | 6 (1.7%) |
| Induration | 5 (2.9%) | 1 (0.6%) | 6 (1.7%) |
| **Subjects with solicited systemic AE** |  |  |  |
| Any | 5 (2.9) | 4 (2.3) | 9 (2.6) |
| General discomfort | 1 (0.6%) | 3 (1.7%) | 4 (1.1%) |
| Fever ($\geq 38^{\circ}\text{C}$) | 2 (1.1) | 1 (0.6) | 3 (0.9) |
| Low-grade fever ($< 38^{\circ}\text{C}$) | 4 (2.3) | 1 (0.6) | 5 (1.4) |

Data are n (%) unless otherwise specified. AE=Adverse Event

Before vaccination, 97.1 % of children were negative for anti-RBD antibodies, median anti-RBD IgG was 1.95 UA/mL (25^th^-75^th^ percentile 1.95; 1.95). Two doses of SOBERANA 02 induced seroconversion in 96.2% participants (CI 95% 93.5; 98.0), satisfying the trial hypothesis (>50% of seroconversion with a lower boundary of the two-sided 95% confidence interval >0.3) (Table 4). By age subgroup, seroconversion was 99.4% (CI 95% 96.5; 99.9) in children 3-11 y/o and 93.1% (CI 95% 88.0; 96.5) in 12-18 y/o. Global seroconversion index was 27.8 (42.6 in children 3-11 y/o, 24.8 in 12-18 y/o); the median anti-RBD IgG was 57.0 UA/mL (25^th^-75^th^ percentile 29.8; 153.4), being higher in younger children (93.3 UA/mL; 25^th^-75^th^ percentile 39.0; 214.4). Specific antibody response was higher than the elicited by natural infection, evaluated in Cuban children convalescent panel (anti-RBD IgG median 11.5; 25^th^-75^th^ percentile 5.3; 24.2). The heterologous third dose increased seroconversion to 100% and seroconversion index to 154.5; anti-RBD IgG titers also increased significantly (p<0.005) to 325.7 UA/mL (25^th^-75^th^ percentile 141.5; 613.8) (Table 4).

**Table 4:** Humoral immune response induced after two doses of SOBERANA 02 and a third heterologous doses with SOBERANA Plus.

| Age groups |  |  |  |  |  |  |  | CCCSP |
| --- | --- | --- | --- | --- | --- | --- | --- | --- |
|  |  | 3-11 years |  | 12-18 years |  | Total |  |  |
|  |  | Post-2<sup>nd</sup> dose | Post-3<sup>rd</sup> dose | Post-2<sup>nd</sup> dose | Post-3<sup>rd</sup> dose | Post-2<sup>nd</sup> dose | Post-3<sup>rd</sup> dose |  |
|  | N | 159 | 156 | 159 | 150 | 318 | 306 | 82 |
| Anti-RBD IgG seroconversion rate | N (%) | 157/158 (99.4) | 155/155 (100.0) | 148/159 (93.1) | 150/150 (100%)* | 305/317 (96.2) | 305/305 (100.0)* | N.D |
|  | 95% CI | 96.5; 99.9 | 99.6; 100.0 | 88.0; 96.5 | 99.6; 100.0 | 93.5; 98.0 | 99.8; 100.0 |  |
| Anti-RBD IgG AU/mL | Median | 93.3 | 329.8* | 49.9 | 325.6* | 57.0 | 325.7* | 11.5 |
|  | 25<sup>th</sup>-75<sup>th</sup> | 39.0; 214.4 | 162.9; 685.6 | 22.3; 90.7 | 108.0; 555.8 | 29.8; 153.4 | 141.5; 613.8 | 5.3; 24.2 |
| Seroconversion index | Median | 42.6 | 155.4* | 24.8 | 154.3* | 27.8 | 154.5* | N.D |
|  | 25<sup>th</sup>-75<sup>th</sup> | 19.5; 97.0 | 75.4; 260.2 | 11.0; 44.7 | 50.2; 266.3 | 14.3; 69.0 | 67.2; 260.9 |  |
| RBD:hACE2 Inh% | Median | 78.3 | 92.5* | 60.1 | 92.2* | 67.4 | 92.4* | 20.8 |
|  | 25<sup>th</sup>-75<sup>th</sup> | 49.4; 88.9 | 88.8; 93.4 | 36.7; 79.7 | 87.7; 93.6 | 42.1; 86.9 | 88.3; 93.5 | 10.9; 40.8 |
| mVNT<sub>50</sub> | GMT | 274.1 | 1418.3* | 143.7 | 1116.2* | 198.5 | 1261.2* | 35.2 |
|  | 95% CI | 216.8; 346.5 | 1180.0; 1704.7 | 115.0; 179.5 | 924.0; 1348.3 | 168.4; 233.9 | 1105.5; 1438.8 | 25.3; 48.9 |
| cVNT<sub>50</sub> vs D614G | N | 63 | 66 | 60 | 65 | 123 | 131 | 70 |
|  | GMT | 28.4 | 181.6* | 24.4 | 137.9* | 26.4 | 158.4* | 9.2 |
|  | 95% CI | 18.5; 43.6 | 120.6; 273.3 | 17.6; 40.0 | 101.8; 186.9 | 20.2; 34.5 | 123.0; 204.0 | 6.8; 12.5 |

T0 or baseline anti-RBD IgG 1.95 (25<sup>th</sup>-75<sup>th</sup> percentile: 1.95; 1.95) for both age subgroups and total. ND: Not determined
Anti-RBD IgG seroconversion rate: % of subjects with seroconversion (CI 95%).
Seroconversion index: increase of IgG concentration respect to baseline (median; 25<sup>th</sup>-75<sup>th</sup> percentile)
AU/mL=anti-RBD IgG concentration expressed in arbitrary units/mL.
RBD:hACE2 Inh%: RBD:hACE2 inhibition % at a serum dilution 1/100.
mVNT<sub>50</sub>: molecular virus neutralization titer; as serum dilution inhibiting 50% of RBD:hACE2 interaction.
cVNT<sub>50</sub>: conventional live-virus neutralization titer.
GMT=Geometric Mean Titer. 95% CI= 95% Confidence Interval; 25<sup>th</sup>-75<sup>th</sup>=25-75 percentile.
\* p<0.005 versus Post - 2<sup>nd</sup> dose McNemar test (anti-RBD IgG seroconversion %), Wilcoxon Signed Ranks test (anti-RBD IgG AU/mL, RBD:hACE2 Inh%) or Paired Student t test (mVNT<sub>50</sub>, cVNT<sub>50</sub>, log-transformed). CCCSP=Cuban children convalescent serum panel

The capacity of anti-RBD IgG for blocking RBD-hACE2 interaction after two doses of SOBERANA 02 was 67.4 % (25^th^-75^th^ percentile 42.1; 86.9) and the mVNT_50_ was 198.5 (CI 95% 168.4; 233.9); both increased significantly (p<0.005) after the third dose to 92.4 % (25^th^-75^th^ percentile 88.3; 93.5) and 1261 (CI 95% 1105,5; 1438.8) respectively. These values were higher among the youngers (3-11 y/o) after the second dose, but were similar in both age subgroups after the third dose. After both two and three doses mVNT50 was higher than after natural infection (Table 4).

After two doses of SOBERANA 02 the neutralizing titer versus D614G variant was higher (GMT 26.4 CI 95% 20.2; 34.5) than the children convalescent panel value (GMT 9.2 CI 95% 6.8; 12.5); and the third dose significantly (p<0.005) boosted the response to GMT 158.4 (CI 95% 123.0; 204.0). The neutralizing titer versus the variants alpha, beta and delta was evaluated in 48 children; 100% had neutralizing antibodies vs. alpha and delta; 97.9% vs. beta (Table 5). GMT was 173.8 (CI 95% 131.7; 229.5) vs. alpha, 142 (CI 95% 101.3; 198.9) vs. delta and 24.8 (CI 95% 16.8; 36.6) vs. beta (a 2.2-fold decrease for delta and 7.0-fold decrease for beta compared to D614G).

**Table 5.** Conventional live-virus neutralization titers against SARS-CoV-2 variants alpha, delta and beta.

|  |  | D614G | Alpha | Delta | Beta |
| --- | --- | --- | --- | --- | --- |
| cVNT<sub>50</sub> | N | 48 | 48 | 48 | 48 |
|  | GMT | 173.8 | 142.0 | 76.8* | 24.8* |
|  | 95% CI | 131.7; 229.5 | 101.3; 198.9 | 54.8; 107.7 | 16.8; 36.6 |

Sera from 48 children vaccinated with complete schedule (two doses SOBERANA 02 + one dose SOBERANA Plus, 28 days apart) were evaluated against D614G, alpha, delta and beta variants. cVNT<sub>50</sub>=conventional live-virus neutralization titer. GMT=Geometric Mean Titer. 95% CI= 95% Confidence Interval. \* p<0.005 Paired Student t test (cVNT<sub>50</sub>, log-transformed) respect to D614G variant.

There was a good correlation among all humoral immunological variables, with coefficients >0.7 (Supplemental Material X-Table S6). Predictive cut-off for attaining cVNT_50_ over 50 was estimated by ROC curve as: 192.2 AU/ml for IgG concentration, 87.1% for the inhibition of RBD:hACE2 and 427 for mVNT_50_ (Supplemental Material X-Table S7, Figure S1)

RBD-specific T cell response in a subset of 45 participants fully vaccinated was determined by measuring IFN-γ and IL-4 expression in peripheral blood mononuclear cells. The number of IFN-γ and IL-4 secreting cells was statistically higher (p<0.001) than their baseline levels (Figure 2), suggesting a mixed Th1/Th2 profile. IFN-γ expression was higher in older children while IL-4 was higher in younger children, evidencing differences in the balance Th1/Th2.

**Figure 2.**
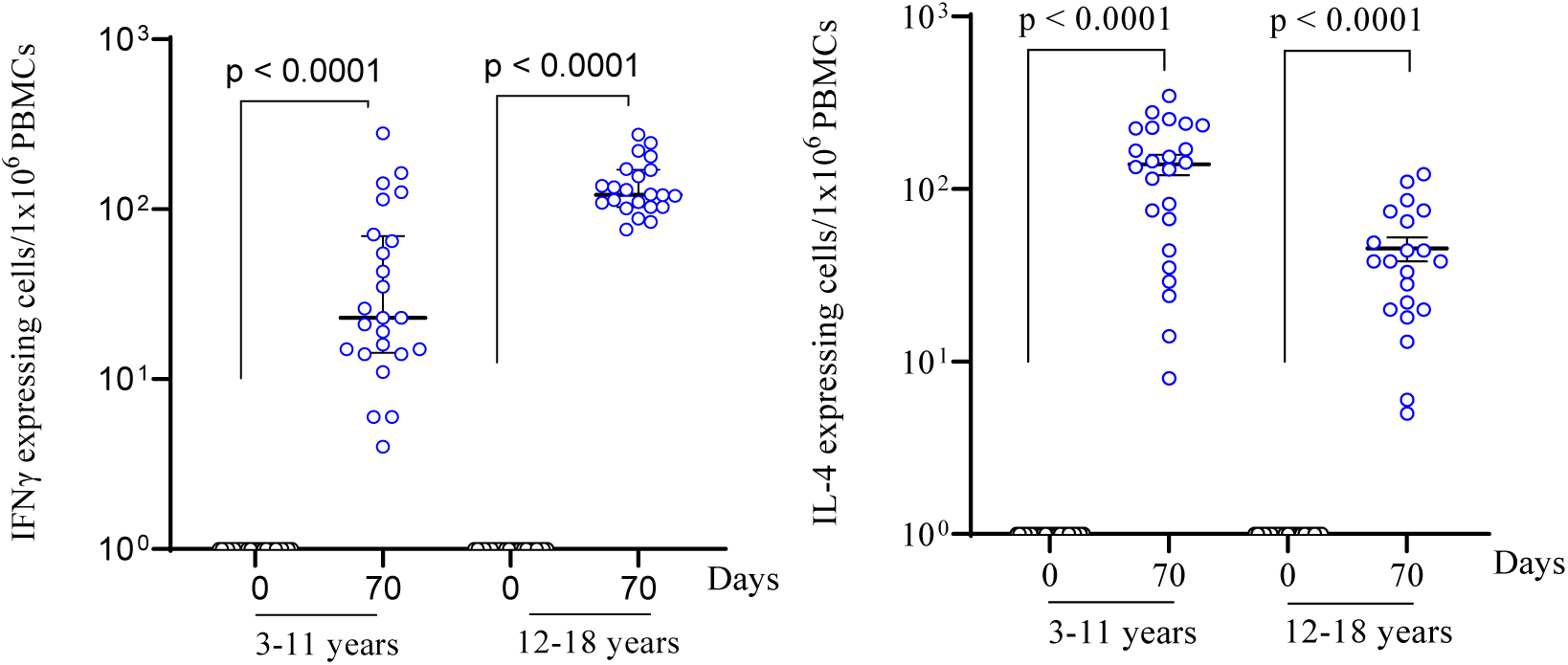
IFN-γ- and IL-4-secreting cells in peripheral blood mononuclear cells stimulated with RBD. Children 3-11 (N=24) and 12-18 years old (N=21) received two doses (on days 0, 28) of SOBERANA 02 and a heterologous third dose (on day 56) of SOBERANA Plus. p value represents the statistic differences as indicated.

The safety and immune response in children were compared with all young adults (aged 19-39) vaccinated in phase I and phase II studies with the same vaccines, as recommended by FDA, 2021; safety profile was similar in both (Supplemental Material X-Table S8, S9, Figure S2). An immunobridging analysis was performed for anti-RBD IgG, mVNT_50_ and cVNT_50_ between children and young adults. IgG elicited after two doses of SOBERANA 02 was 57.0 UA/ml (25^th^-75^th^ percentile 29.8; 153.4), while for young adults it was 46.4 (25^th^-75^th^ percentile 17.4; 108.8); after the heterologous third dose of SOBERANA Plus these values increased to 325.7 (25^th^-75^th^ percentile 141.5; 613.8) in children and 228.0 (25^th^-75^th^ percentile 95.8; 394.3) in young adults. mVNT_50_ was 198.5 (IC 95% 168.4; 233.9) in children after the second dose and 1261.2 (IC 95% 1105.5; 1438.8) after the third; in young adults were 94.9 (IC 95% 75.0; 120.2) and 503.7 (IC 95% 432.6; 586.6) after two and three doses (Figure 3). We found significant differences (p<0.05) for IgG and mVNT_50_ between 3-18 y/o children and 19-39 y/o young adults; higher values were obtained in children after both the second and the third dose. Viral neutralization titers after the second dose were measured at different time points in children and young adults (on day 42 in children and day 56 in young adults), making their comparison only approximate.

**Figure 3:**
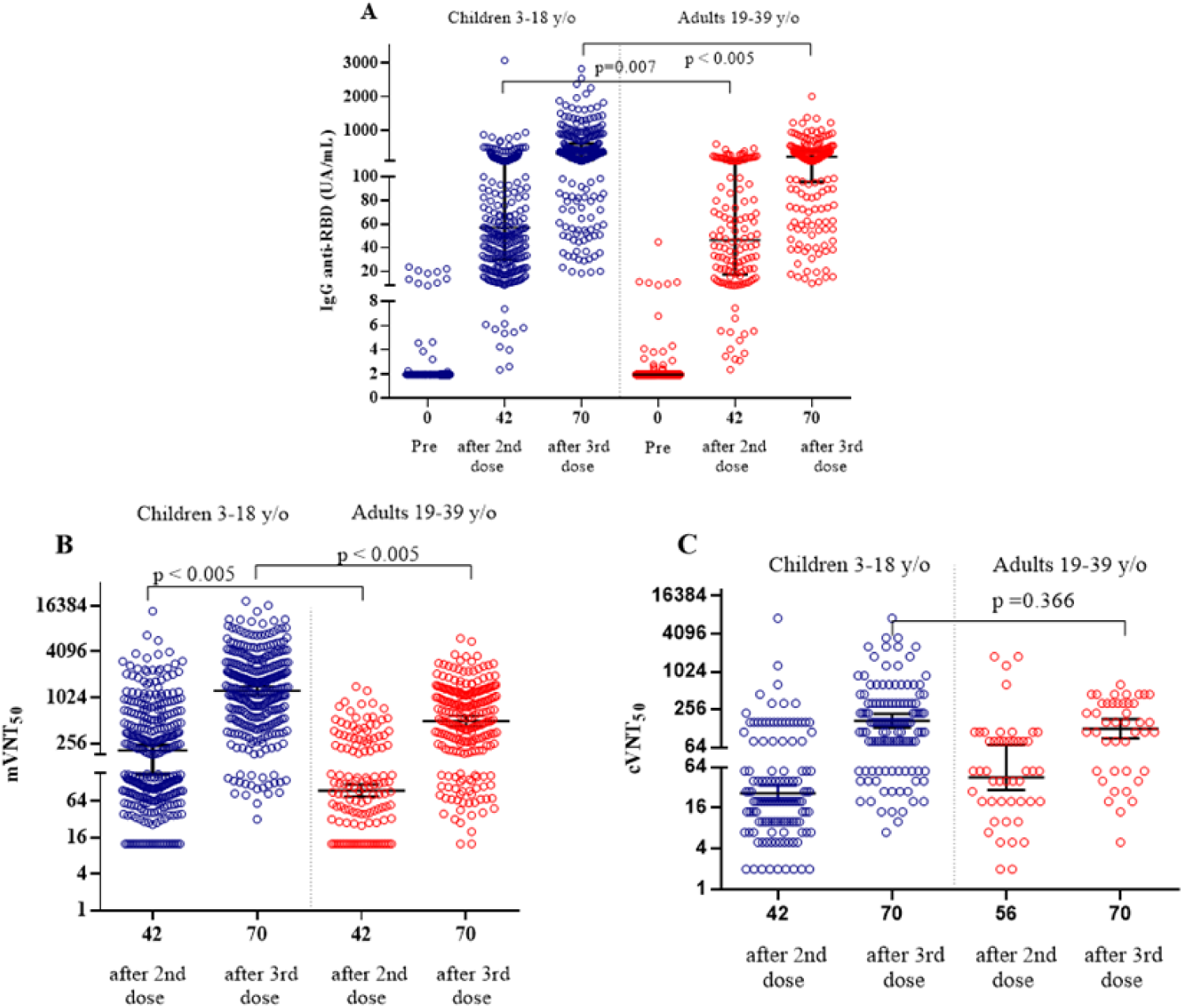
Immunobridging comparison of humoral immune response elicited in children (3-18 y/o) respect to young adults (19-39 y/o from Phase I and II clinical trial), after two doses of SOBERANA 02 (day 42) and a third dose of SOBERANA Plus (day 70). A) anti-RBD IgG median (25^th^-75^th^ percentile); B) mVNT_50_ GMT (CI 95%); C) cVNT_50_ GMT (CI 95%). Bleeding was on day 42 and 70 (14 days after second or third dose), except for cVNT50 adults after second dose was on day 56. Mann-Whitney U test (anti-RBD IgG AU/mL) or Student t test (mVNT_50_, cVNT_50_, log-transformed).

The non-inferiority analysis was performed with cVNT_50_ data, following FDA recommendation (FDA, 2021). After three doses (on day 70), cVNT_50_ in children was 158.4 (IC 95% 123.0; 204.0) and 122.8 (80.2; 188.0) for young adults (n=43, data available) (Figure 3). The immune response in 3-18 y/o, as well as in age subgroups 3-11 y/o and 12-18 y/o was non-inferior to that observed in 19-39 y/o young adults. The geometric mean ratio (3-18 y/o value to young adult’s value) for cVNT_50_ 14 days after the third dose was 1.25 (CI 95% 0.77; 2.02) (Table 6), which met the non-inferiority criterion (i.e., a lower boundary of the two-sided 95% confidence interval of >0.67). In summary, both age groups (3-11 y/o and 12-18 y/o) met the non-inferiority criterion.

**Table 6:**
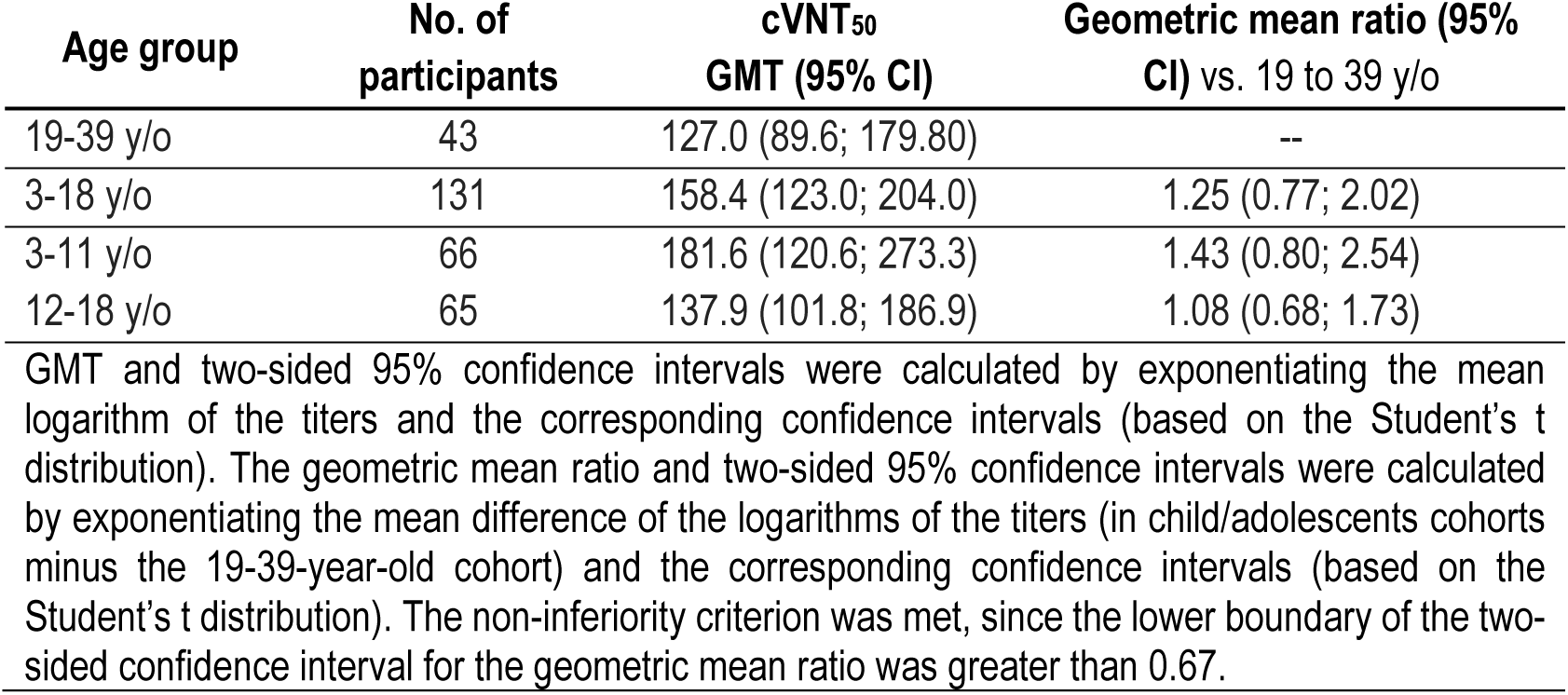
Immunobridging of cVNT_50_ in children and young adults after heterologous scheme (two doses of SOBERANA 02 and a third heterologous doses of SOBERANA Plus)

Based on immunogenicity data of vaccinated children and the immune response to natural infection (children convalescent panel), a prediction of clinical efficacy was estimated through a regression linear model. By using cVNT_50_ as the predictive variable, the estimated efficacy is 91.3% (CI 95% 84.6; 95.1) after two doses and 97.4 % (IC 95% 91.5; 99.2) after three doses (Figure 4). This study was conducted in Havana during delta wave; after receiving the first dose 29 children (8.2%) of 350 children became PCR-positive, this value dropped down to 13 (4.0%) out of 321 children after completing the two-dose scheme.

**Figure 4.**
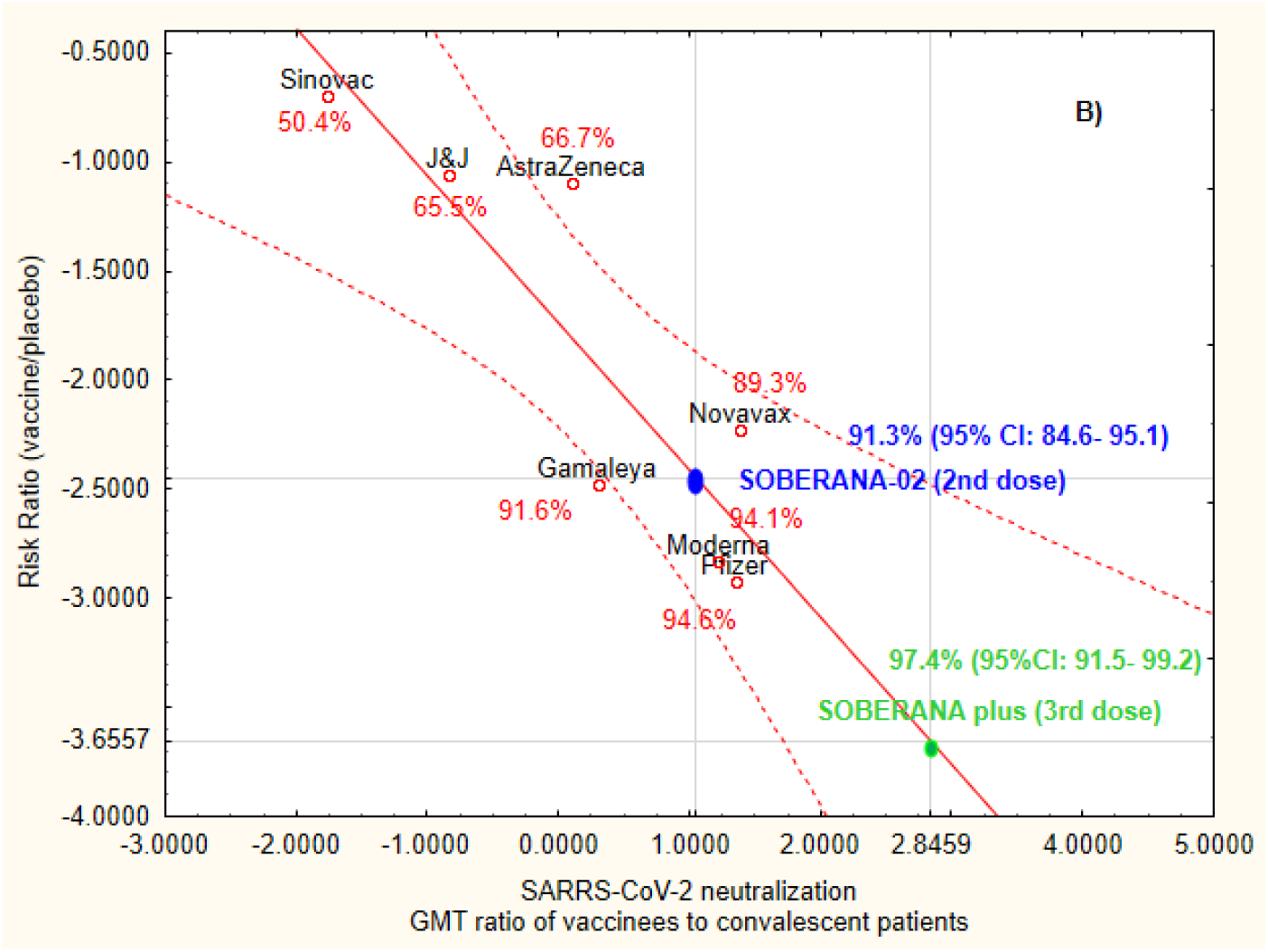
Prediction of clinical efficacy in children from correlation between antibody responses and efficacy rate. Panels displays correlation of cVNT_50_ neutralization and ratios, respectively for seven vaccines in adults; SOBERANA 02 and SOBERANA Plus in children. The y-axis is estimated log risk ratio reported on the vaccine efficacy scale. The x-axis is log ratio of the peak geometric mean neutralization at 7-28 days post-vaccination, relative to human or children convalescent sera.

## Discussion

This study describes, for a first time, the safety and immunogenicity in children 3-18 y/o of two doses of SOBERANA 02, followed by a heterologous third doses of SOBERANA Plus. The frequency of local and systemic AE was 49.0% and 2.6% respectively, lower than after mRNA COVID-19 vaccination (Frenck et al., 2021; Ali et al., 2021). After two doses, BNT162b2 reported 86.0% and 66.0 of children 12-15 y/o with local and systemic AEs while mRNA-1273 reported 94.2% and 68.3% (aged 12-17 y/o) respectively (Frenck et al., 2021; Ali et al., 2021). In our study, local pain was reported by the 51.4% of children aged 12-18 y/o after first dose, 17% after the second and 17.3% after the third one. BNT162b2 and mRNA-1273 vaccine in adolescents reported 86.0% and 94.2% with local pain after first dose; and 79.0% and 92.4% after the second, respectively (Frenck et al., 2021; Ali et al., 2021). SOBERANA 02 and SOBERANA Plus caused general discomfort (the most frequent systemic AE) only in 1.7% of children 12-18 y/o, while mRNA vaccines provoked fatigue, headache, chills, muscle pain or fever in 10-68.5% of adolescents (Frenck et al., 2021; Ali et al., 2021). In children aged 3-11 local pain was the unique EA with frequency >10% during this study; children aged 5-11 vaccinated with BNT162b2 reported local pain (74.0%), redness (19.0%), swelling (15.0%), fatigue (39.0%) and headache (28.0%) (Walter et al., 2021). Myocarditis and pericarditis have been reported in adolescent after mRNA COVID-19 vaccination (Oster et al., 2021; Marshall et al., 2021); these AEs were not observed here.

The comparison of the humoral immune response elicited by vaccination with the elicited by natural infection has been a useful tool for the development of several anti-SARS-CoV-2 vaccine (Keech et al., 2020; Yang et al., 2021). Two shots of SOBERANA 02 every 28 days in children induced a robust humoral response, with higher levels of antibodies and similar neutralizing capacity of the elicited by natural infection. The third dose of SOBERANA Plus boosted both the production of antibodies and their neutralizing capacity, surpassing the immune response in convalescent children, as had been previously observed in clinical trials in adults (Toledo-Romani et al., 2021a; 2021b).

The induction of specific T-cell response is critical for protection of viral infections. The heterologous three doses schedule in children developed a balanced activation of IFN-γ and IL-4-secreting cells from PBMC, indicating a mixed Th1/Th2 response, as reported in adults after the same vaccination scheme (Toledo-Romani et al., 2021a).

The SARS-CoV-2 variants of concern alpha, beta, delta, and recently, omicron have modified the pandemic landscape worldwide (Fontane et al., 2021). Here, we report the capacity of anti-RBD antibodies for neutralizing alpha, beta and delta variant, with a fold-reduction of 2.2 for delta and 7.0 for beta compared to D614G, as we found in adults (Toledo-Romani et al., 2021b). In an independent study from the “Pedro Kourí” Tropical Medicine Institute in Havana, sera from 20 adults vaccinated with SOBERANA 02 + SOBERANA Plus neutralize the omicron variant (Portal-Miranda 2022; Carles JM, 2022). Recently, the Pasteur Institute of Iran informed that Pasteurcovac, the Iranian brand of SOBERANA 02, is 100% effective against the omicron variant in adults (Biglari 2022).

We conducted this clinical trial during the delta wave, the worst period of Cuban epidemic (Rodriguez; 2021); in such a context and due to ethical reasons, a placebo-controlled clinical trial was not ethical. Lacking a control group (main limitation of the trial), two analytical tools complemented the study: immunobridging with the immune response in young adults previously vaccinated during clinical trial with same vaccination schedule (no concurrent reference population) as recommended by FDA, 2021; and prediction of clinical efficacy based on immunological response (Khoury et al., 20211; Kristen et al., 2021). Firstly, we found a non-inferior response for the ratio of SARS-CoV-2 cVNT_50_ after the three-dose scheme in participants 3-11 and 12-18 y/o relative to a 19-39 y/o reference population (no concurrent) The comparison met the non-inferiority criterion with ratio 1.43 (CI 95% 0.8-2.54) for 3-11 y/o and 1.08 (CI 95% 0.68-1.73) for 12-18 y/o, satisfying FDA recommendations (FDA, 2021). Similar analyses have been reported by BNT162b2 and mRNA-1273 vaccines but using 19-25 y/o as reference population (Walter et al., 2022; Ali et al., 2021). Based on published results, we considered young adults as immunocompetent up to 39 years (Lopez-Sejas 2016; Ventura et al., 2017; Thapa and Farber 2019); this increased the number of cVNT_50_ data for comparison in the reference population. Secondly, a prediction of clinical efficacy based on immunological response has been advanced for other vaccines (Khoury et al., 20211; Kristen et al., 2021). Using this model, for adults aged 19-80 we anticipated a clinical efficacy between 58% and 87% after first two doses and between 81% and 93% after the three-dose scheme (Toledo-Romani et al., 2021b). These results were confirmed during a phase III clinical trial reporting a 71% of efficacy for the two-dose schedule of SOBERANA 02 and 92.4% for the heterologous three-dose schedule (Toledo-Romani et al., 2021c). Here, the model predicts 89.3%-91.3% clinical efficacy after two doses of SOBERANA 02 and 95.1%-97.4% after the third doses of SOBERANA Plus in children versus the D614G strain.

Starting children vaccination at 2 y/o is key for controlling the pandemic, cutting the transmission and reducing the emergence of new VOCs (Petersen and Buchy, 2021). The safety and immunological results reported here supported the emergency use authorization of SOBERANA 02 and SOBERANA Plus as heterologous scheme for children between 2-18 y/o. A massive immunization campaign started on 5^th^ September 2021; fully vaccinating 1.8 millions of Cuban children (96% of 2-18 y/o Cuban population (Redd, 2022; Augustin, 2022). These results support public health vaccination strategies, providing children as young as two years a safe and effective vaccine scheme to prevent COVID-19.

## Data Availability

All data produced in the present study are available upon reasonable request to the authors

https://www.finlay.edu.cu/blog/category/sala-cientifica/

## Declaration of Interests

The authors R.P.G, Y.R.D, C.R.I, L.C.H. M.P.B, D.V.M, N.P.P, C.V.S, A.P.D, E.G.O, Y.C.E, J.C.F.P, M.S.M, M.D.E, J.M.E.P, Y.I.H, M.E.T.R declare that they have no known competing financial interests or personal relationships that could have appeared to influence the work reported in this paper.

The authors B.P.M, M.R.G, B.S.R, L.M.R.N, R.P.N, R.G.M, T.H.G, T.F.B, I.C.Q, S.F.C, Y.C.R, D.S.M, Y.G.V, Y.V.B, D.G.R, V.V.B work at Finlay Vaccine Institute or the Centre of Molecular Immunology, institutions that develop and manufacture the vaccine candidates but they haven’t received an honorarium for this paper.

B.S.R., S.F.C., Y.C.R, L.R.N., D.S.M., Y.V.B., D.G.R. and V.V.B., have filed patent applications related to the vaccine SOBERANA 02.

D.D and A.B work at Pasteur Institute of Iran and are co-developer of the vaccines

## Acknowledgments

We especially thanks all the parents and children for participated in the clinical trial. We recognize the contribution of all the medical and nurse staff at clinical site. We thank Dr. Lila Castellanos for scientific advice.

