## Supplementary material for "Open label phase I/II clinical trial and predicted efficacy of SARS-CoV-2 RBD protein vaccines SOBERANA 02 and SOBERANA Plus in children": Suplemmental material

**SUPPLEMENTAL MATERIAL**

**INDEX**

I. **Trial Sites**
II. **Selection criteria.**
III. **Products under evaluation**
IV. **Outcomes.**
V. **Safety assessment**
VI. **Immunogenicity assessment and techniques**
VII. **Cuban children convalescent serum panel (CCCSP)**
VIII. **Statistical analysis.**

**IX. Supplementary safety data.**

**X. Supplementary immunogenicity data.**

I. **Trial Sites**

Phase I was conducted at “Juan Manuel Marquez” Pediatric Hospital; phase II was conducted also at policlinics “5 de Septiembre” and “Carlos J. Finlay” all sites in Havana, Cuba. Clinical sites were certified by the National Immunization Program

II. **Selection criteria.**

**II.1 Inclusion criteria:**

1. Subjects aged 3-18 y/o .

2. Voluntariness expressed through informed consent to participate in the study:

- Subjects 3-11 y/o : Informed consent of parents or legal guardians

- Subjects 12-18 y/o : Informed Consent of the parents or legal guardians and informed assent of the adolescent.

3. Weight-height nutritional assessment in the 10^th^ - 90^th^ percentile (for subjects 3- 9 y/o ); Body Mass Index in the 10^th^ - 90^th^ percentile for subjects 10- 18 y/o ), according to the standardized nutritional measurement for the Cuban pediatric population.

4. Physical examination: normal or without clinically significant alterations

5. For potential participants in phase I: clinical laboratory results within range of reference values, or outside but without clinically significance (only).

**II.2. Exclusion criteria:**

1. Acute febrile or infectious disease at the time of the vaccine application or in the 7 days prior to product administration.

2. History of SARS-CoV-2 and COVID-19 who meet any of the following criteria: a) Previous or current history of SARS-CoV-2 infection. b) Be declared in the category of contact or suspect at the time of inclusion. c) Subject with positive test for anti-SARS-CoV-2 antibodies. d) Subject with positive PCR at the time of inclusion.

13. Participation in another clinical trial in the last 3 months.

14. Application of vaccines containing tetanus toxoid in the last 3 months.

15. Application of other vaccines in the last 30 days.

3. History of hypersensitivity to thiomersal or any of the components of the formulations.

4. History of having been immunized with a SARS-CoV 2 vaccine.

5. History of having received a vaccine from the Cuban immunization scheme, in a period of≤ 30 days before product administration.

6. Use of any product under investigation in ≤ 30 days before immunization.

7. Application of vaccines containing tetanus toxoid in the last 3 months.

8. History of chronic diseases.

9. History of major congenital malformations (defects that have a significant functional compromise for the individual's life, have medical consequences and require early, sometimes urgent, care).

10. Primary or secondary immune system disease.

11. History of neoplastic disease.

12. History of severe allergic reactions.

13. Treatment with immunomodulators in the last 30 days (i.e., steroids (except topical and inhaled), Interferon, Immunoferon, Nasalferon, Transfer Factor, monoclonal antibody, Biomodulin T, any gammaglobulin, Heberferon, Thymosin, Levamisole).

14. Subjects with a history of convulsive disease.

15. History of treatment with blood products such as blood cells, plasma, whole blood or platelet concentrate transfusions in the last 4 months.

16. Splenectomy or splenic dysfunction.

17. Child with a minor or mentally disabled mother or father.

18. Pregnancy or lactation (a pregnancy test will be carried out before inclusion and administration of each dose to all girls and adolescents who menstruate).

19. Subjects with tattoos in the deltoid region of both arms.

20. Subjects with a history or positive results for: antibodies against HIV1 + 2, antibodies against hepatitis C, surface antigen of the hepatitis B virus or VDRL serology.

21. History of psychoactive substance use in the last 6 months.

III. **Products under evaluation**

**Table S1. Composition of vaccines**

| **Ingredient** | **Vaccines** | |
| --- | --- | --- |
|  | **SOBERANA 02** | **SOBERANA Plus** |
| Antigen | SARS-CoV-2 RBD conjugated to  tetanus toxoid, 25 µg RBD  per 20 µg tetanus toxoid | SARS-CoV-2 RBD dimer  (d-RBD), 50 µg |
| Aluminium hydroxide | 0.5 mg | 1.25 mg |
| Sodium chloride | 4.25 mg | 4.25 mg |
| Disodium hydrogen phosphate | 0.03 mg | 0.03 mg |
| Sodium dihydrogen phosphate | 0.02 mg | 0.02 mg |
| Water for injection | 0.5 ml | 0.5 ml |

IV. **Outcomes.**

IV.1 Primary outcomes

- *Primary outcome for Phase I*: Serious Adverse Events (SAE)
  - a) Occurrence of the SAE (Yes, No),
  - b) Duration (time since the beginning until the end of the event);
  - c) Description of the event;
  - d) Result (recovered, recovered with squeals, persists, death, unknown);
  - e) Causality association (consistent causal relation to immunization, Inconsistent causal relation to immunization, Indeterminate, Unclassifiable)
  - Clinical evaluation: daily for 28 days after each dose.
- *Primary Outcome for phase II:* Concentration of specific anti-RBD IgG antibodies and percentage of subjects with ≥4-fold increase in antibody titers over pre-immunization titers. Evaluation on samples collected on day 0, 42 and 70.

IV.2 Secondary outcomes

*- Solicited local and systemic adverse events (AE); (both phases)*

- Occurrence of the AE (Yes, No)
- Duration (days since the beginning until the end of the event)
- Intensity of the AE (mild, moderate, severe)
- Severity (serious, not serious)
- Result (recovered, recovered with sequelae, persists, death, unknown)
- Causality (causal association consistent with vaccination, undetermined, causal association inconsistent with vaccination, not classifiable)
- Data collection: daily, for 7 days after each dose.

*- Unsolicited Adverse Events (AE); (both phases)*

- Occurrence of the AE (Yes, No)
- Duration (days since the beginning until the end of the event)
- Intensity of the AE (mild, moderate, severe)
- Severity (serious, not serious)
- Result (recovered, recovered with sequelae, persists, death, unknown)
- Causality (causal association consistent with vaccination, undetermined, causal association inconsistent with vaccination, not classifiable)
- Data collection: daily, for 28 days after each dose.
- *Serious Adverse Events (SAEs) (phase II)*
  - a) Occurrence of the SAE (Yes, No),
  - b) Duration (time since the beginning until the end of the event);
  - c) Description of the event;
  - d) Result (recovered, recovered with squeals, persists, death, unknown);
  - e) Causality association (consistent causal relation to immunization, Inconsistent causal relation to immunization, Indeterminate, Unclassifiable)
  - Clinical evaluation: daily for 28 days after each dose.
- *Conventional live-virus neutralization titer* *cVNT_50_* *(both phases)*: Evaluation on samples collected on days 42 and 70 versus D614G variant, for a subset of subjects with seroconversion. Evaluation on samples collected on day 70 (for a subset of subjects) versus alpha, beta and delta variant
- *% RBD-hACE2-inhibition*: *(both phases):* Evaluation on samples collected on day 0 (only for positive IgG samples), 42, 70.
- *Molecular virus neutralization titer* *mVNT_50:_ (both phases):* Evaluation on samples collected on day 0 (only for positive IgG samples), 42, 70.
- *RBD-specific T-cells responses producing IFN- γ and TNF- α*: *(phase II):* Evaluation on samples collected on days 0, 70.

V. **Safety assessment**

Pain, erythema, swelling, induration and temperature were local-solicited AEs at the injection site. Fever ≥38 ^◦^C, low-grade fever (<38 ^◦^C), general discomfort and rash were systemic-solicited AEs. Any other events were parent-recorded throughout the 28 days follow-up period after each dose. Clinical laboratory test included pre-vaccination and post-vaccination biochemical serum analysis (only during phase I).

AEs were classified as serious or not serious; severity—according to Brighton Collaboration definition and the Common Terminology Criteria for AE version 5·0—was: mild when AE was transient (not interfering with activities), moderate (when caused mild to moderate limitation in activity), or severe (limitating activity). AEs were reviewed for causality and classified according to WHO: inconsistent causal association to immunization, consistent causal association to immunization, undetermined, unclassifiable (WHO, 2018).

VI. **Immunogenicity assessment and techniques**

*VI.1. Anti-RBD IgG response*

Anti-RBD IgG in sera was evaluated on days 0, 42 and 70, by a quantitative ultramicro ELISA (UMELISA SARS-CoV-2 anti- RBD, Centre for Immunoassay, Havana, Cuba) using d-RBD as coating antigen (4 µg/mL) and an in-house standard characterized serum, which was arbitrarily assigned 200 AU/mL (based on a half-maximal inhibitory titer of 200 and a conventional virus neutralization titer of 160). The standard curve comprised two-fold serial dilutions (0, 4, 8, 16, 32 and 64 AU/mL) of the standard. Samples were evaluated in duplicate. After incubation , biotin-conjugate anti-IgG human (0.1 µg/mL) (Sigma Aldrich, San Luis, EE UU) and then, streptavidin/alkaline-phosphatase (Roche, Basel, Swiss) in appropriate buffer were added. The final fluorimetric reaction was induced by adding the substrate 4-Methylumbelliferyl Phosphate (Sigma Aldrich, San Luis, EE UU). The reference curve was constructed using a linear interpolation function. The concentration of anti-RBD IgG was expressed as AU/mL; negative samples were reported as 1.95 AU/mL; samples over 7.8 AU/mL were positive. The seroconversion rate was calculated by dividing the concentration at each time point (at Tx) by the pre-vaccination concentration (at T0). A rate ≥ 4 was considered as seroconversion.

*VI.2 Inhibitory capacity of antibodies for blocking the RBD-hACE2 interaction*

A competitive ELISA determined the inhibitory capacity of antibodies for blocking the RBD-hACE2 interaction. It was expressed as % inhibition and molecular virus neutralization titer (mVNT_50_) and was evaluated on days 0 (only for positive IgG samples), 42 and 70. Microtiter plates were coated with 250ng/well of ACE2-hFc in carbonate-bicarbonate buffer, 0.1M (pH9.6) and incubated overnight at 4°C. Plates were blocked with 200µL/well of 2% of skim milk in PBST (PBS with Tween 20 0.05%) during 1 h, at 37°C. Serial dilutions of sera were pre-incubated with RBD-mouse-Fc (RBD-mFc) at a final concentration of 20 ng/mL, for 1 h at 37^o^C. These mixtures were added to the plates and incubated for 2 h at 37°C. The binding of RBD-mFc was detected by addition of alkaline phosphatase-conjugated anti-mouse IgG antibody (Sigma) for 1 h at 37°C. Finally, p-nitrophenylphosphate (Sigma) at 1 mg/mL in diethanolamine buffer (pH9.8) was added, and plates were incubated at RT for 30 min. The OD at 405nm was measured using a microwell system reader (BioTek). In all steps other than blockade, samples and reagents were added to a final volume of 50 µL/well. Three washing steps with PBST followed all incubations. RBD-mFc, sera and antibody conjugates were diluted in skim milk 0.2%/PBST. Inhibition was expressed as percentage according to the formula: Inhibition (%)= [1-(OD_405nm_sample/OD_405nm_ maximal recognition)] x 100. Maximal recognition corresponds to wells incubated only with RBD-mFc (20 ng/mL). For determination of mVNT_50_, dilutions were log transformed and the highest dilution giving 50% of inhibition was calculated.

*VI.3. Conventional Virus Neutralization titer*

This test—the gold standard for determining antibody efficacy against SARS-CoV-2 (Manenti et al; 2020)— using live SARS-CoV-2 was performed in a biosecurity laboratory level 3 (National Civil Defense Research Laboratory, Havana, Cuba) by the conventional virus neutralization test,. Conventional virus neutralization titer (cVNT_50_) vs. D614G strain was evaluated in a subset of samples randomly selected from the individuals with seroconversion after two doses (on day 42, n=123) and after three doses (on day 70, n=131). Another subset of samples (n=48) was randomly selected for neutralizing tests vs alpha, beta, and delta variants after the third dose (on day 70). Serial dilutions of heat-inactivated serum samples (starting from 1:5) in Eagle’s Minimal Essential Medium (Gibco, UK) containing 2 % fetal bovine serum (Capricorn, Germany) were incubated for 1 hour at 37°C with an equal volume of viral solution containing 100 TCID_50_ of SARS-CoV-2 (Strains: CU2010-2025, variant D614G; CU2101-2102, variant B.1.1.7 alpha; CU2104-2179, variant B.1.617.2 delta; CU2104-2180, variant B.1.351 beta; Cuban Collection at National Civil Defence Research Laboratory) in cell plates containing a semiconfluent VeroE6 monolayer (10^4^ cell/well). The highest serum dilution, showing an OD at 540 nm representing the 50% of average OD values from control cell wells (VeroE6 monolayer with mixture of virus-serum) was considered as the neutralization titer and is represented as conventional virus neutralization titer 50 (cVNT_50_).

*VI.4 Specific T-cell response*

RBD-specific T-cell response producing IFN- γ and IL-4 were quantified by enzyme-linked immunospot (ELISpot) assay using human IFN-γ ELISpot^PLUS^ HRP kit (Mabtech, Sweden) and human IL-4 ELISpot^plus^ HRP kit (Mabtech, Sweden) following the manufacturer’s instructions. Peripheral blood mononuclear cells (PBMCs) were isolated on day 0 (before first immunization) and 70 (14 days after the third dose) from a subset of children 3-11 (N=24) and 12-18 y/o (N=21). RBD-specific T-cell response producing IFN- γ and IL-4 were quantified with enzyme-linked immunospot (ELISpot) assay using human IFN-γ ELISpot^PLUS^ HRP kit (Mabtech, Sweden) and human IL-4 ELISpot^plus^ HRP kit (Mabtech, Sweden) following the manufacturer´s instructions. Briefly, cryopreserved peripheral blood mononuclear cells from vaccinated subjects were thawed and rested for 6 hours in RPMI 1640 medium supplemented with 1000 units/mL penicillin, 1 mg/mL streptomycin, 1 mM pyruvate (Gibco), 50 µM β-mercaptoethanol (Sigma-Aldrich) and 10% *(v/v)* heat-inactivated fetal calf serum (Capricorn). PBMCs (2.5 x 10^5^ cells/well, in duplicate) were stimulated with recombinant RBD (10 µg/mL) at 37°C for 24 h before detection. Specific T-cell response was expressed as the number of spot-forming cells per 10^6^ cells; measurements were subtracted from the unstimulated control values.

VII. **Cuban children convalescent serum panel (CCCSP)**

A Cuban children convalescent serum panel (CCCSP) was made with sera from 82 patients (3-18 y/o) recovered from COVID-19: five from children with moderate symptomatic disease, 29 with mild disease and 48 asymptomatic; concerning prevalent variant of concern, 21 from delta wave and 61 from pre-delta wave when D614G and beta variants predominated in Havana. Convalescent children were attended by specialized doctors; their parents gave written consent for studying their immunological status after natural infection allowing their use for epidemiological research. This panel was characterized by anti-RBD IgG concentration (UA/ml), inhibition of RBD-hACE2 interaction (% of inhibition and molecular neutralization titer) and virus neutralization titer (cVNT_50_) as described in Supplementary Material VI.

VIII. **Statistical analysis.**

Safety and reactogenicity endpoints are described as frequencies (%). Quantitative demographic characteristics are reported as mean, standard deviation (SD), median, interquartile range, and range. Seroconversion rate for anti-RBD IgG antibodies (≥4-fold increase in antibody concentration over baseline) was expressed as % with the 95% confidence interval and seroconversion index as median and interquartile range. Anti-RBD IgG concentration and % of inhibition of RBD-hACE2 interaction were expressed as median and interquartile range; mVNT50 and cVNT50 were expressed as geometric mean titer (GMT) and 95% confidence intervals (CI).

Spearman´s rank correlation was used to assess relationships among techniques used to evaluate the immune response. The cut—off value (for IgG concentration, % inhibition RBD:hACE2 and mVNT_50_) for predicting cVNT_50_ over 50 was established according to ROC curves,. The Student t-Test or the Wilcoxon Signed-Rank Test were used for before-after statistical comparison.

A stopping criterion due to unacceptable toxicity (frequency >1%) was evaluated iteratively, with a Bayesian algorithm. As an additional analysis, a non-inferiority of the immune response in 3-11 and 12-18 y/o participants compared with that in 19-39 y/o participants (from not concurrent phase I and phase II studies (Toledo-Romani et al., 2021a; 2021b) was assessed, using the geometric mean ratio of SARS-CoV-2 50% neutralizing titers (cVNT_50_). The geometric mean ratio and two-sided 95% confidence intervals were calculated by exponentiating the mean difference of the logarithms of the titers and the corresponding confidence intervals (based on the Student’s t distribution). The non-inferiority criterion was met, since the lower boundary of the two-sided confidence interval for the geometric mean ratio was >0.67 (FDA, 2021).

A prediction of clinical efficacy was estimated using the regression linear model reported for seven vaccines based on immunogenicity data (Khoury et al 20211; Earle et al., 2021). Statistical analyses were done using SPSS version 25.0; STATISTICA version 12.0, R version 3.2.4, EPIDAT version 3.1 and Prism GraphPad version 6.0. An alpha signification level of 0.05 was used.

**IX. Supplementary safety data.**

**Table S2.** Global characterization of adverse events

|  | **Age groups** | |  |
| --- | --- | --- | --- |
|  | **3-11 years** | **12-18 years** | **Total** |
| **Total adverse events** | 141 | 182 | 323 |
| **Intensity** |  |  |  |
| Mild (Grade 1) | 135 (95.7) | 167 (91.8) | 302 (93.5) |
| Moderate (Grade 2) | 6 (4.3) | 15 (8.2) | 21 (6.5) |
| Severe (Grade 3) | 0 | 0 | 0 |
| **Severity** |  |  |  |
| No Serious | 141 (100.0) | 181 (99.5) | 322 (99.7) |
| Serious | 0 | 1 (0.5) * | 1 (0.3) * |
| **Causal relationship**** |  |  |  |
| Consistent (A1) | 126 (89.4) | 160 (87.9) | 286 (88.5) |
| Consistent (A2) | 0 | 0 | 0 |
| Consistent (A3) | 0 | 0 | 0 |
| Indeterminate (B1) | 0 | 2 (1.1) | 2 (0.6) |
| Indeterminate (B2) | 0 | 1 (0.5) | 1 (0.3) |
| Inconsistent | 15 (10.6) | 19 (10.4) | 34 (10.5) |
| **Result** |  |  |  |
| Recovered | 140 (99.3) | 180 (98.9) | 320 (99.1) |
| Persist | 0 | 1 (0.5)* | 1 (0.3)* |
| **Type** |  |  |  |
| Local | 118 (83.7) | 147 (80.8) | 265 (82.0) |
| Systemic | 23 (16.3) | 35 (19.2) | 58 (18.0) |
| **Solicited** |  |  |  |
| Solicited | 125 (88.7) | 152 (83.5) | 277 (85.8) |
| Unsolicited | 16 (11.3) | 30 (16.5) | 46 (14.2) |
| **Starting at** |  |  |  |
| ≤ 60 min | 25 (13.7) | 25 (17.7) | 50 (15.5) |
| 60 min-24 hours | 127 (69.8) | 85 (60.3) | 212 (65.6) |
| 24-48 hours | 15 (8.2) | 14 (9.9) | 29 (9.0) |
| 48-72 hours | 1 (0.5) | 3 (2.1) | 4 (1.2) |
| > 72 hours | 14 (7.7) | 14 (9.9) | 28 (8.7) |
| **Duration (hours)** |  |  |  |
| ≤ 24 hours | 99 (54.4) | 74 (52.5) | 173 (53.6) |
| 24-48 hours | 37 (20.3) | 35 (24.8) | 72 (22.3) |
| 48-72 hours | 33 (18.1) | 16 (11.3) | 49 (15.2) |
| > 72 hours | 13 (7.1) | 16 (11.3) | 29 (9.0) |
| Footnote: Frequency: number of events, %: values referred to total events.  *** Dengue.  *** A1:* Vaccine-related event (according to published literature)*; A2:* Event related to a defect in the quality of the vaccine*; A3:* Error-related reaction*; A4:* Event related to the conditions inherent to the vaccinated subject*; B1:* The temporal relationship is consistent, but the evidence is insufficient to consider the vaccination as cause of the event*; B2:* Classification criteria result in contradiction regarding consistencies and inconsistencies with a causal association with immunization | | | |

.

**Table S3**. Frequency of subjects with adverse event by dose

|  |  | **Age groups** | | |
| --- | --- | --- | --- | --- |
|  |  | **3-11 years** | **12-18 years** | **Total** |
| **N** |  | 175 | 175 | 350 |
| **Overall adverse events within 28 days after vaccination** | | | | |
| **Subject with AE** | | 81 (46.3) | 105 (60.0) | 186 (53.1) |
| 1^st^ dose | | 54/175 (30.9) | 98/175 (56.0) | 152/350 (43.4) |
| 2^nd^ dose | | 29/162 (17.9) | 29/159 (18.2) | 58/321 (18.1) |
| 3^rd^ dose | | 30/156 (19.2) | 31/150 (20.7) | 61/306 (19.9) |
| **Subjects with solicited AE within 7 days after vaccination** | | | | |
| **Any** | | 76 (43.4) | 98 (56.0) | 174 (49.7) |
| 1^st^ dose | | 50/175 (28.6) | 90/175 (51.4) | 140/350 (40) |
| 2^nd^ dose | | 27/162 (16.7) | 27/159 (17.0) | 54/321 (16.8) |
| 3^rd^ dose | | 29/156 (18.6) | 28/150 (18.7) | 57/306 (18.6) |
| **Subjects with solicited local adverse events** | | | | |
| **Any** | | 74 (42.3) | 98 (56.0) | 172 (49.1) |
| 1^st^ dose | | 50/175 (28.6) | 90/175 (51.4) | 140/350 (40) |
| 2^nd^ dose | | 27/162 (16.7) | 27/159 (17.0) | 54/321 (16.8) |
| 3^rd^ dose | | 26/156 (16.7) | 27/150 (18.0) | 53/306 (17.3) |
| **Local pain** | | 69 (39.4) | 98 (56.0) | 167 (47.7) |
| 1^st^ dose | | 49/175 (28) | 90/175 (51.4) | 139/350 (39.7) |
| 2^nd^ dose | | 23/162 (14.2) | 27/159 (17.0) | 50/321 (15.6) |
| 3^rd^ dose | | 19/156 (12.2) | 26/150 (17.3) | 45/306 (14.7) |
| **Swelling** | | 9 (5.1) | 2 (1.1) | 11 (3.1) |
| 1^st^ dose | | 2/175 (1.1) | 0/175 (0.0) | 2/350 (0.6) |
| 2^nd^ dose | | 3/162 (1.9) | 0/159 (0.0) | 3/321 (0.9) |
| 3^rd^ dose | | 6/156 (3.8) | 2/150 (1.3) | 8/306 (2.6) |
| **Local warm** | | 4 (2.3) | 0 (0.0) | 4 (1.1) |
| 1^st^ dose | | 0/175 (0.0) | 0/175 (0.0) | 0/350 (0.0) |
| 2^nd^ dose | | 2/162 (1.2) | 0/159 (0.0) | 2/321 (0.6) |
| 3^rd^ dose | | 2/156 (1.3) | 0/150 (0.0) | 2/306 (0.7) |
| **Erythema** | | 5 (2.3) | 1 (0.6) | 6 (1.7) |
| 1^st^ dose | | 0/175 (0.0) | 0/175 (0.0) | 0/350 (0.0) |
| 2^nd^ dose | | 2/162 (1.2) | 0/159 (0.0) | 2/321 (0.6) |
| 3^rd^ dose | | 4/156 (2.6) | 1/150 (0.7) | 5/306 (1.6) |
| **Induration** | | 5 (2.3) | 1 (0.6) | 6 (1.7) |
| 1^st^ dose | | 2/175 (1.1) | 0/175 (0.0) | 2/350 (0.6) |
| 2^nd^ dose | | 1/162 (0.6) | 0/159 (0.0) | 1/321 (0.3) |
| 3^rd^ dose | | 3/156 (1.9) | 1/150 (0.7) | 4/306 (1.3) |
| **Subjects with solicited systemic adverse events** | | | | |
| **Any** | | 5 (2.9) | 4 (2.3) | 9 (2.6) |
| 1^st^ dose | | 1/175 (0.6) | 3/175 (1.7) | 4/350 (1.1) |
| 2^nd^ dose | | 0/162 (0.0) | 0/159 (0.0) | 0/321 (0.0) |
| 3^rd^ dose | | 4/156 (2.6) | 1/150 (0.7) | 5/306 (1.6) |
| **Mild fever** | | 4 (2.3) | 1 (0.6) | 5 (1.4) |
| 1^st^ dose | | 0/175 (0.0) | 1/175 (0.6) | 1/350 (0.3) |
| 2^nd^ dose | | 0/162 (0.0) | 0/159 (0.0) | 0/321 (0.0) |
| 3^rd^ dose | | 4/156 (2.6) | 0/150 (0.0) | 4/306 (1.3) |
| **Fever** | | 2 (1.1) | 1 (0.6) | 3 (0.9) |
| 1^st^ dose | | 1/175 (0.6) | 0/175 (0.0) | 1/350 (0.3) |
| 2^nd^ dose | | 0/162 (0.0) | 0/159 (0.6) | 0/321 (0.3) |
| 3^rd^ dose | | 1/156 (0.1) | 1/150 (0.7) | 2/306 (0.7) |
| **General discomfort** | | 1 (0.6) | 3 (1.7) | 4 (1.1) |
| 1^st^ dose | | 0/175 (0.0) | 3/175 (1.7) | 3/350 (0.9) |
| 2^nd^ dose | | 0/162 (0.0) | 0/159 (0.0) | 0/321 (0.0) |
| 3^rd^ dose | | 1/156 (0.6) | 0/150 (0.0) | 1/306 (0.3) |

**Table S4**. Unsolicited adverse events.

|  | **Age groups** | | |
| --- | --- | --- | --- |
|  | **3-11 years** | **12-18 years** | **Total** |
| **N (%)** | 175 (100.0%) | 175 (100.0%) | 350 (100.0%) |
| Number of participants reporting unsolicited AEs | 14 (8.0) | 21 (12.0) | 35 (10.0) |
| Vaccine-related | 5 (2.9)* | 10 (5.7)** | 15(4.3) |
| Headache | 2 (1.1) | 9 (5.1) | 11 (3.1) |
| Non Vaccine-related | 9 (5.1)*** | 13 (7.4)**** | 22 (6.3) |
| Headache | 1 (0.6) | 2 (1.1) | 3 (0.9) |
| Footnote: *Reported by 1 subject (0.6%, each one): arm cramp, drowsiness, heavy arm sensation.  ** Reported by 1 subject (0.6%, each one): arm cramp  *** Reported by 1 subject (0.6%, each one): diarrhea; gastrointestinal disease; epistaxis; boil in ear; foot wound; viral process; nasal secretion; cough  **** Reporting by 1 subject (0.6%, each one): cellulitis on thigh, tonsillitis; miliaria; dengue; diarrhea; fever; mild fever, hypertension; upper acute respiratory infection; abdominal discomfort; nasal secretion; cough. Reporting by 2 subjects (1.1% each one): vomiting; sickness, diarrhea | | | |

**Table S5. Haematology and blood chemistry evaluation for children (only phase I)**

|  | | | **3-11 years** | **12-18 years** | **Global** |
| --- | --- | --- | --- | --- | --- |
| N | | | 25 | 24 | 49 |
| **Haemoglobin** | Reference values (g/L) | | 115-135 | F: 120-160; M: 130-160 |  |
|  | Pre | Media (SD) | 130.5 (8.4) | 138.6 (13.7) | 134.9 (10.3) |
|  |  | (Min; Max) | (116;154) | (117;162) | (113;165) |
|  | Day 7 | Media (SD) | 132.1 (6.8) | 137.9 (12.4) | 134.5 (11.9) |
|  |  | (Min; Max) | (117;148) | (113;165) | (116;162) |
|  | **p vs. 0 (t Student)** | | 0.127 | 0.719 | 0.667 |
|  | Day 70 | Media (SD) | 127.9 (6.3) | 137.8 (14.2) | 132.6 (11.8) |
|  |  | (Min; Max) | (119; 143) | (116; 167) | (116; 167) |
|  | **p vs. 0 (t Student)** | | 0.130 | 0.807 | 0.435 |
| **Hematocrit** | Reference values | | 2-6 years: 0.34-0.40  7-11years: 0.37-0.49 | F: 0.35-0.45  M: 0.37-0.49 |  |
|  | Pre | Media (SD) | 0.36 (0.02) | 0.38 (0.03) | 0.37 (0.03) |
|  |  | (Min; Max) | (0.32; 0.42) | (0.32; 0.44) | (0.32; 0.44) |
|  | Day 7 | Media (SD) | 0.36 (0.02) | 0.39 (0.03) | 0.37 (0.03) |
|  |  | (Min; Max) | (0.31; 0.40) | (0.32; 0.46) | (0.31; 0.46) |
|  | **p vs. 0 (t Student)** | | 0.041 | 0.957 | 0.326 |
|  | Day 70 | Media (SD) | 0.35 (0.02) | 0.38 (0.04) | 0.37 (0.03) |
|  |  | (Min; Max) | (0.33; 0.39) | (0.33; 0.46) | (0.33; 0.46) |
|  | **p vs. 0 (t Student)** | | 0.039 | 0.979 | 0.344 |
| **Platelets** | Reference values (10^9^/L) | | 150-450 | | |
|  | Pre | Media (SD) | 280 (62) | 231 (51) | 256 (61) |
|  |  | (Min; Max) | (175; 410) | (161; 323) | (161; 410) |
|  | Day 7 | Media (SD) | 288 (54) | 253 (60) | 271 (59) |
|  |  | (Min; Max) | (197; 409) | (160; 380) | (160; 409) |
|  | **p vs. 0 (t Student)** | | 0.355 | 0.178 | 0.099 |
|  | Day 70 | Media (SD) | 280 (55) | 221 (36) | 252 (55) |
|  |  | (Min; Max) | (175; 389) | (169; 285) | (169; 389) |
|  | **p vs. 0 (t Student)** | | 0.649 | 0.514 | 0.885 |
| **Leukocytes** | Reference values (10^9^/L) | | 2-6 years: 5.0-14.5  7-11 years: 4.5-13.5 | 4.5 – 11.0 |  |
|  | Pre | Mediana (IRI) | 7.2 (1.8) | 5.5 (1.9) | 6.3 (2.1) |
|  |  | (Min; Max) | (4.9; 11.3) | (0.3; 8.6) | (0.3; 11.3) |
|  | Day 7 | Mediana (IR) | 7.2 (2.4) | 7.2 (1.5) | 7.2 (2.2) |
|  |  | (Min; Max) | (4.7; 10.8) | (6.0; 9.0) | (4.7; 10.8) |
|  | **p vs. 0 (Wilcoxon)** | | 0.162 | 0.000 | 0.000 |
|  | Day 70 | Mediana (IR) | 7.3 (2.0) | 6.8 (1.6) | 7.0 (1.8) |
|  |  | (Min; Max) | (3.7; 12.3) | (3.7; 9.7) | (3.7; 12.3) |
|  | **p vs. 0 (Wilcoxon)** | | 0.648 | 0.006 | 0.013 |
| **Glucose** | Reference values (mmol/L) | | 3.88 -6.1 | | |
|  | Pre | Media (SD) | 4.2 (0.3) | 4.7 (0.4) | 4.4 (0.4) |
|  |  | (Min; Max) | (3.4; 4.7) | (4.0; 5.7) | (3.4; 5.7) |
|  | Day 7 | Media (SD) | 4.3 (0.5) | 4.2 (0.3) | 4.3 (0.4) |
|  |  | (Min; Max) | (3.3; 5.3) | (3.6; 4.8) | (3.2; 5.3) |
|  | **p vs. 0 (t Student)** | | 0.103 | 0.000 | 0.025 |
|  | Day 70 | Media (SD) | 3.4 (0.4) | 4.2 (0.4) | 3.8 (0.5) |
|  |  | (Min; Max) | (2.7; 4.1) | (2.9; 5.2) | (2.7; 5.2) |
|  | **p vs. 0 (t Student)** | | 0.000 | 0.000 | 0.000 |
|  | Reference values (μmol/L) | | 27-65 μmol/L | 46-77 μmol/L |  |
| **Creatinine** | Pre | Media (SD) | 47.9 (7.8) | 69.2 (14.6) | 58.3 (15.8) |
|  |  | (Min; Max) | (30.0; 62.0) | (48.1; 96.3) | (30.0; 96.3) |
|  | Day 7 | Media (SD) | 43.6 (6.8) | 52.6 (18.8) | 48.0 (14.6) |
|  |  | (Min; Max) | (31.0; 53.0) | (18.5; 82.9) | (18.5; 82.9) |
|  | **p vs. 0 (t Student)** | | 0.000 | 0.000 | 0.000 |
|  | Day 70 | Media (SD) | 45.2 (7.7) | 68.3 (14.6) | 56.3 (16.3) |
|  |  | (Min; Max) | (26.0; 58.0) | (39.0; 92.0) | (26.0; 92.0) |
|  | **p vs. 0 (t Student)** | | 0.037 | 0.854 | 0.106 |
| **ASAT** | Reference values (U/L) | | Up to 49 U/L | | |
|  | Pre | Media (SD) | 32.3 (5.9) | 22.0 (3.5) | 27.2 (7.1) |
|  |  | (Min; Max) | (22.0; 46.0) | (16.3; 29.4) | (16.3; 46.0) |
|  | Day 7 | Media (SD) | 21.8 (6.9) | 18.6 (2.0) | 20.2 (5.3) |
|  |  | (Min; Max) | (12.0; 36.0) | (15.6; 23.6) | (12.0; 36.0) |
|  | **p vs. 0 (t Student)** | | 0.000 | 0.000 | 0.000 |
|  | Day 70 | Media (SD) | 27.8 (4.9) | 17.4 (3.8) | 22.8 (6.8) |
|  |  | (Min; Max) | (19.0; 38.0) | (12.0; 27.0) | (12.0; 38.0) |
|  | **p vs. 0 (t Student)** | | 0.001 | 0.000 | 0.000 |
| **ALAT** | Reference values (U/L) | | Up to 49 U/L | | |
|  | Pre | Median (IR) | 21.0 (5.4) | 12.2 (4.3) | 18.0 (9.8) |
|  |  | (Min; Max) | (15.0; 41.0) | (7.6; 34.4) | (7.6; 41.0) |
|  | Day 7 | Median (IR) | 17.0 (9.0) | 12.2 (5.3) | 13.4 (7.3) |
|  |  | (Min; Max) | (10.0; 53.0) | (7.6; 29.4) | (7.6; 53.0) |
|  | **p vs. 0 (Wilcoxon)** | | 0.034 | 0.897 | 0.027 |
|  | Day 70 | Median (IR) | 17.9 (9.5) | 14.3 (8.4) | 16.7 (9.1) |
|  |  | (Min; Max) | (9.0; 57.0) | (7.0; 37.0) | (7.0; 57.0) |
|  | **p vs. 0 (Wilcoxon)** | | 0.010 | 0.948 | 0.027 |

**X. Supplementary immunogenicity data.**

**Table S6.** Correlations between immunological variables after two doses of SOBERANA 02 and a third dose of SOBERANA Plus.

|  | | | **Anti-RBD IgG (UA/mL)** | **% Inh RBD:hACE2** | **mVNT_50_** |
| --- | --- | --- | --- | --- | --- |
| **After 2 doses** | **Anti-RBD IgG AU/mL** | r^2^ Spearman | 1.000 |  |  |
|  |  | p | . |  |  |
|  |  | N | 318 |  |  |
|  | **% Inh RBD:hACE2** | r^2^ Spearman | 0.892** | 1.000 |  |
|  |  | p | 0.000 | . |  |
|  |  | N | 318 | 318 |  |
|  | **mVNT_50_** | r^2^ Spearman | 0.909** | 0.970** | 1.000 |
|  |  | p | .000 | .000 | . |
|  |  | N | 318 | 318 | 318 |
|  | **cVNT_50_** | r^2^ Spearman | 0.881** | 0.885** | 0.898** |
|  |  | p | 0.000 | 0.000 | 0.000 |
|  |  | N | 123 | 123 | 123 |
| **After 3 doses** | **Anti-RBD IgG UA/mL** | r^2^ Spearman | 1.000 |  |  |
|  |  | p | . |  |  |
|  |  | N | 306 |  |  |
|  | **% Inh RBD:hACE2** | r^2^ Spearman | 0.715** | 1.000 |  |
|  |  | p | 0.000 | . |  |
|  |  | N | 306 | 306 |  |
|  | **mVNT_50_** | r^2^ Spearman | 0.859** | 0.827** | 1.000 |
|  |  | p | 0.000 | 0.000 | . |
|  |  | N | 306 | 306 | 306 |
|  | **cVNT_50_** | r^2^ Spearman | 0.804** | 0.729** | 0.874** |
|  |  | p | 0.000 | 0.000 | 0.000 |
|  |  | N | 131 | 131 | 131 |

Footnotes: After two doses: Two doses of SOBERANA 02 every 28 days; After three doses: Two doses of SOBERANA 02 and a third dose with SOBERANA Plus every 28 days. ** p<0.01

**Table S7.** Predictive value of immunological variable for attaining conventional live-virus neutralization titer >50 (cVNT_50_ titer>50).

| Test Result Variable(s) | Cutt-off | Area | Std. Error | Sig. | 95% Confidence Interval | |
| --- | --- | --- | --- | --- | --- | --- |
|  |  |  |  |  | Lower Bound | Upper Bound |
| Anti-RBD IgG (AU/mL) | 192.2 | 0.916 | 0.028 | 0.000 | 0.860 | 0.971 |
| % Inh RBD:hACE2 | 87.1 | 0.960 | 0.016 | 0.000 | 0.928 | 0.992 |
| mVNT_50_ | 427.5 | 0.952 | 0.018 | 0.000 | 0.917 | 0.987 |

**Figure S1.** ROC Curve for cVNT_50_ ≥ 50

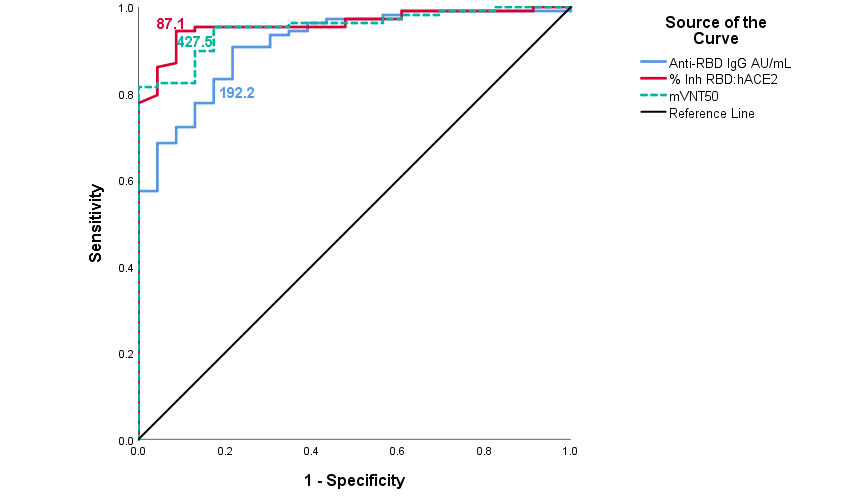

**VII. Supplementary data for immunobridging subset.**

**Table S8**. Demographic characteristics of the subjects included in immunobridging subset

|  |  | **Children**  **3-18 years** | **Young adults groups as reference**  **19 -39 years** |
| --- | --- | --- | --- |
| **N** |  | 131 | 43 |
| **Sex** | |  |  |
| Female | | 60 (45.8) | 27 (62.8) |
| Male | | 71 (54.2) | 16 (37.2) |
| **Skin colour** | |  |  |
| White | | 84 (64.1) | 30 (69.8) |
| Black | | 9 (6.9) | 4 (9.3) |
| Multiracial | | 38 (29.0) | 9 (20.9) |
| **Age** | |  |  |
| Mean (SD) | | 11.2 (4.6) | 28.3 (5.9) |
| Median (IR) | | 11.0 (8.0) | 28.0 (10.0) |
| Range | | 3-18 | 19-39 |

**Table S9**. Subjects 3-18-years-old and 19-39-years-old reporting at least 1 adverse event

|  | **Age groups** | |
| --- | --- | --- |
|  | **3-18 years** | **19-39 years** |
| **N** | 350 | 261 |
| Subjects with some AE | 186 (53.10%) | 148 (56.7%) |
| Subjects with some VAAE | 177 (50.6%) | 133 (51.0%) |
| Subjects with some Serious AE | 1 (0.3%)* | 3 (1.1%)** |
| Subjects with some Serious VAAE | 0 (0.0%) | 0 (0.0%) |
| Subjects with some Severe AE | 0 (0.0%) | 1 (0.4%)*** |
| Subjects with some Severe VAAE | 0 (0.0%) | 0 (0.0%) |
| * Dengue  ** 1 subject with: Lipothymia- Rectal bleeding-Diarrhea; 1 subject with upper acute respiratory infection; 1 subject with bilateral orchiepididymitis  *** bilateral orchiepididymitis | | |

**Figure S2**. Comparison of solicited adverse events in all participants in the 3–18 y/o group (N=350) and the subset of young adult 19–39 y/o (N=261)

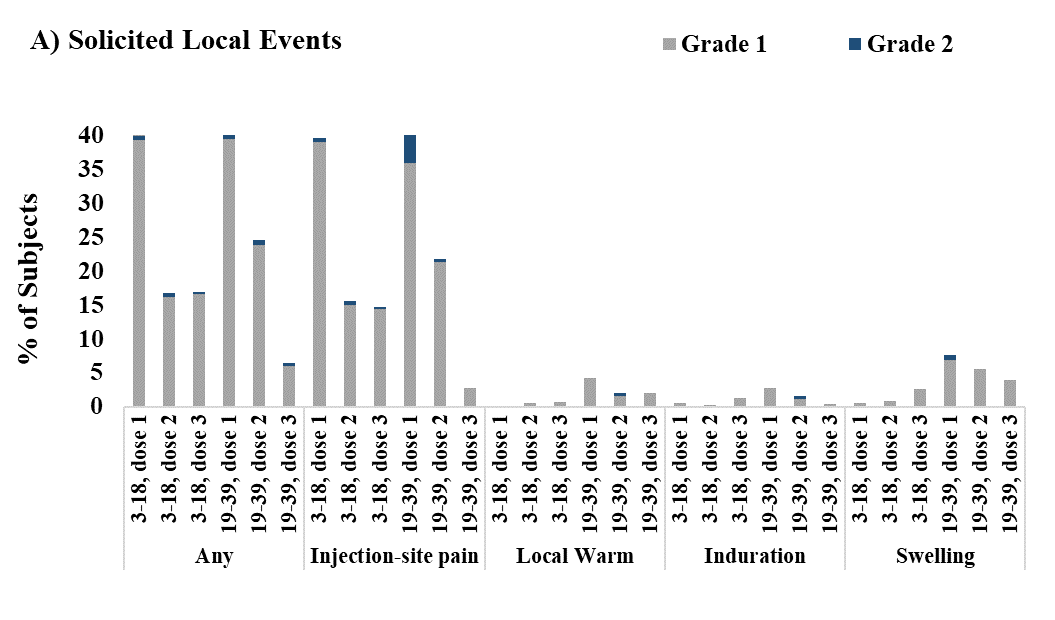

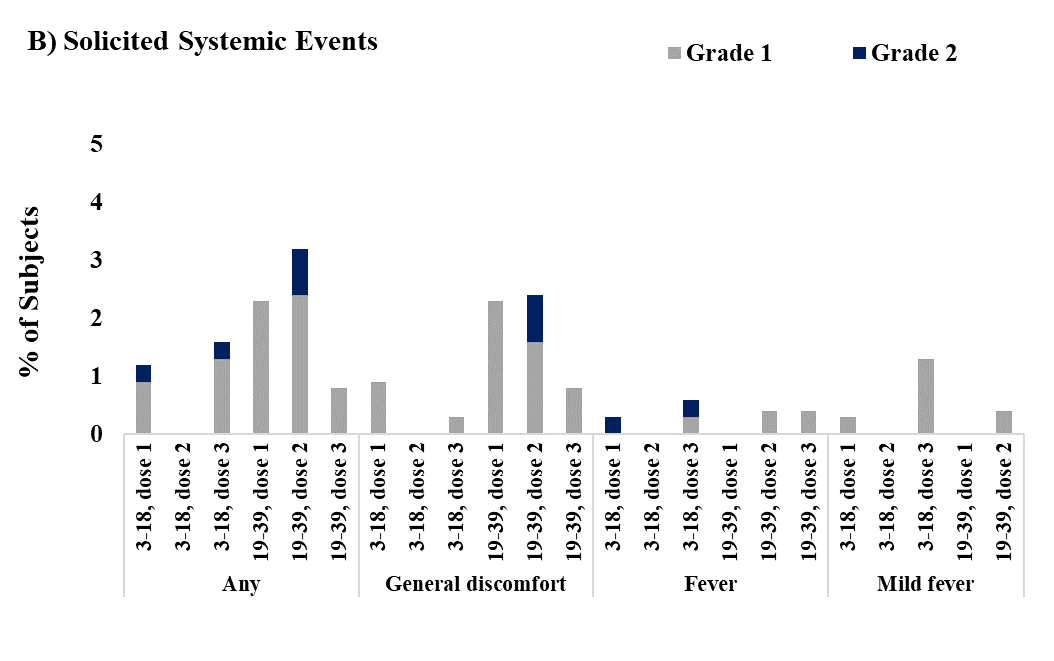

Footnote: Grade 1= Mild; Grade 2= Moderate
